# Epigenetic Effects of HIV and Childhood Maltreatment in Women

**DOI:** 10.64898/2026.09.11.26362833

**Authors:** Aqeedah Abbas Roomaney, Sian Megan Joanna Hemmings, Georgina Spies, Scott Letendre, Soraya Seedat, Jacqueline Samantha Womersley

## Abstract

**Introduction:** People with HIV have disproportionately higher rates of adverse childhood experiences compared to the general population. This, in turn, is linked to long-term physical and mental health consequences. DNA methylation - a heritable, reasonably stable and quantifiable epigenetic mechanism implicated in gene silencing - may offer a useful lens that helps understand how HIV and childhood trauma become biologically embedded and affect disease risk profiles.

**Methods:** This study aimed to identify regional and network level methylomic signatures associated with HIV and childhood maltreatment scores in South African women who bear the highest dual burden. DNA methylation profiling using the Illumina Infinium EPIC V2 kit was performed on DNA extracted from blood samples of 238 women with or without HIV with varying levels of childhood maltreatment. Regional DNA methylation analyses, composed of correlation tests on specific CpG sites and on genetic regions, were conducted for each of HIV and childhood maltreatment scores. Thereafter, a weighted gene Co-Expression Network Analysis (WGCNA) was conducted on HIV and childhood maltreatment scores to determine co-expression modules associated with the variable of interest. Finally, causal mediation analysis of the most significant network module was conducted to clarify the direction of association.

**Results:** Neither HIV status nor childhood maltreatment was associated with DNA methylation at site-specific or regional levels. Network methylation effects were, however, observed, with three out of four network modules that were differentially methylated in HIV, implicating genes involved in immune pathways. One network module, composed of an abundance of genes relating to neurotransmitter functioning, was associated with childhood maltreatment. Both causal analyses implied that HIV and childhood maltreatment preceded the epigenetic effects.

**Conclusions:** The findings suggest that DNA methylation in immune pathways may partially explain the immune and neurologic effects of HIV. The link between childhood maltreatment and neuronal processes warrants further investigation.

## Introduction

More than 40 million people live with HIV[1], with sub-Saharan Africa accounting for about 70% of the global disease burden [2–4]. South Africa is considered a global epicentre of HIV, with an infection prevalence of 13.9%, compared to the global estimate of 0.5 – 0.6% [5].

Women account for 54% of people with HIV (PWH) globally [6]. In South Africa, HIV prevalence is 8% in women older than 14 years and 9.1% in women between the ages of 15 and 24 years, the most affected group [7]. In contrast, the prevalence of HIV amongst South African men 15 years and older is 4.2%, with the prevalence in men between the ages of 15 and 24 years being 3% [7]. In South Africa, an estimated 70% of PWH receive antiretroviral therapy (ART), with 64% of PWH being virally suppressed [8].

### Childhood trauma

Childhood trauma (CT), the experience of emotional, physical, or sexual abuse or physical or emotional neglect of a child [9], has an estimated prevalence of 60-70% in South Africa and may impact long-term physical and psychological health [10–13]. Traumatic experiences may become biologically embedded, impacting neurodevelopment, and creating latent vulnerability to poorer resilience and health [14]. PWH often experience high levels of CT [15], at higher rates than the general population [16,17]. Higher reported CT is associated with poor ART adherence and higher viral loads in PWH, producing a double burden on health and wellbeing [17]. The epigenetic effects of HIV and CT may influence physical and psychological health.

### Epigenetics – DNA methylation

Epigenetic regulation refers to biological processes that regulate gene expression without altering the genetic sequence, and functions as a molecular interface between the genome and the environment [18]. DNA methylation, a reasonably stable and easily quantifiable form of epigenetic regulation in which the addition of a methyl group to a cytosine nucleotide, can influence regional gene expression and ultimately phenotypic changes by regulating transcriptional activity, providing valuable regulatory information [19,20]. Multiple immune response-related genes are susceptible to epigenetic modification [21]. HIV infection affects DNA methylation, particularly expression of immune-regulating genes [22–27]. DNA methylation may also represent a mechanism for the biological embedding of early traumatic experiences such as CT [28], with epigenome-wide DNA methylation studies identifying differentially methylated genes associated with CT [29–32].

### Aim and rationale

Considerable advancements have been made using candidate gene approaches to understand the role of CM and of HIV on DNA methylation. However, candidate gene studies do not account for the complexity of physiological processes, which rely on multiple gene products acting together [28,33]. Examining wider changes in DNA methylation is necessary to better understand how it is affected by HIV and CM. This study investigated the epigenetic profiles of HIV and CM, and their combined effects, in a cohort of South African women (n = 238; women with HIV = 124, women without HIV = 114), using a hypothesis-free approach.

## Methods

Participants were recruited from a parent study at Stellenbosch University, “Biological endophenotypes of HIV and childhood trauma: a genetics, cognitive, and imaging study” [34]. Ethics approval was obtained from the Stellenbosch University Health Research Ethics Committee [N07/07/153]. Biological measures, blood specimens, and neuropsychological data were obtained from the parent study.

### Participant recruitment

In the parent study, women were recruited from community health care facilities in and around Cape Town, South Africa between 2008 and 2010, with a subset were followed over time. Eligibility criteria included age 18 - 65 years, at least a fifth-grade level of literacy in English, Afrikaans, or isiXhosa, and capacity to provide informed consent. Participants with schizophrenia, bipolar disorder, other psychotic disorders, and drug or alcohol abuse at the time of assessment were excluded, as determined on the MINI-International Neuropsychiatric Interview-Plus [35], which was administered by a trained research psychologist. Additional criteria for exclusion included previous head trauma, current seizure disorder, central nervous system infection or psychotropic medication use in the preceding month. Socio-demographic measures such as age, level of education, employment status, and if applicable, ART regimen, and past use of psychotropic medication were recorded.

### Biological measures

Blood was collected for DNA extraction, the assessment of virologic and immune markers, and confirmation of HIV status using enzyme-linked immunosorbent assay. DNA was extracted from blood buffy coat using a phenol-chloroform method or the Qiagen Blood and Tissue DNA extraction kit (Qiagen, Germany) as per the manufacturers protocol. Extracted DNA was stored at –80 °C.

### Childhood maltreatment

Experience of childhood maltreatment (CM) was determined using the Childhood Trauma Questionnaire – Short Form (CTQ-SF) [36], a 28-item self-report measure of the experience of emotional, physical, and sexual abuse, and emotional and physical neglect before the age of eighteen years. Scores for each of the 5-item subscales are combined to generate a total CTQ-SF score ranging from 25 - 125. Higher CTQ-SF scores indicate higher CM, with a score of less than 31 indicating little to no experience. We used the continuous total CTQ-SF score for regional analysis whilst the network analysis utilized a cut-off score of ≥ 31 to indicate the experience of childhood trauma for each participant [37] .

### DNA methylation

DNA extracted from whole blood samples was assessed for quality and purity using the Qubit 4 fluorometer (Invitrogen, Waltham, MA) or the NanodropOne spectrophotometer (ThermoFisher Scientific, Waltham, MA). DNA samples were diluted to a fixed mass of 350 ng total DNA. The DNA, with a normalized concentration of 6 ng/μl, was loaded onto hard-shell 96-well PCR plates (Biorad, Hercules, CA) and sealed using microseal foil seals (Biorad, Hercules, CA), prior to their shipment.

A total of three shipments of extracted DNA samples were sent to the University of Minnesota Genomics Center for bisulphite conversion and DNA methylation profiling, noting the batch number for each sample. Upon reaching the Genomics Center, DNA samples were subjected to a second set of quality assessments, including re-quantification of DNA, followed by an assessment of DNA integrity using quantitative polymerase chain reaction (qPCR). DNA methylation profiling was performed using the Infinium Methylation EPIC v2 Bead Chip kit (Illumina, San Diego, CA), which contains more than 900,000 CpG probes covering the human genome, with high reproducibility and consistency among technical replicates [38].

### DNA methylation quality control

Quality control and normalization of the DNA methylation data was conducted using the R (https://www.r-project.org/) package *Bioconductor: ENmix* [39] and included assessment of multi-density and multi-frequency plots. Four outlier and low-quality samples and 10473 probes were removed. A sample was considered an outlier if its value was more than three times smaller than the lower quartile or three times larger than the upper quartile. As part of the normalization process, background and dye bias correction was performed followed by inter-array normalization, probes-type bias correction, and finally batch effect correction. Cell-type proportion estimates were generated for each sample using the FlowSorted.Blood.EPIC reference data. Cell proportions were calculated for B-cells, CD4T, CD8T, monocyte, neutrophil, and natural killer cells. A surrogate variable analysis was conducted to estimate batch effects and unknown experimental confounders using intensity data for non-negative internal control probes, as outlined in the *ENmix* users guide.

### Covariate selection

A type III ANOVA was conducted to determine drivers of variation in DNA methylation. Based on this, age, batch number, cell type proportions, and three principal components identified in the surrogate variable analysis were included as covariates. Analyses using CM as the predictor variable also included HIV status as a covariate. Due to high missingness in smoking data (36.6%), we were unable to determine the influence of smoking in our analyses. In the subset of participants with smoking data, smoking status did not differ between HIV groups.

### CpG-association analysis

A CpG-association analysis was conducted on DNA methylation data using the R package *cpgassoc* [40]. We first examined associations between DNA methylation and the CTQ-SF total scores and HIV infection separately. Next, we included both CTQ-SF scores and HIV status as independent variables in the analysis. Multiple testing correction was conducted using the Benjamini-Hochberg procedure for all analyses.

### Gene enrichment analysis

Gene enrichment analysis was conducted on the top 100 CpG sites found during the CpG-association analysis. Using the ‘gometh’ function of the R package *missMethyl*, the analysis accounts for two sources of bias: first, the differing number of probes per gene set on the array, and second, CpG sites that map to multiple genes. We tested enrichment using the Gene Ontology (GO) database with genomic features set to include all genomic regions.

### DNA methylation regional (DMR) analysis

A DNA methylation regional analysis was conducted using the Bioconductor R package *DMRcate*, which identifies and ranks the most differentially methylated regions across the genome [41]. First, the individual associations between DMRs and each of HIV status and CTQ-SF scores were tested. Thereafter, CTQ-SF total scores and HIV were included in the same model.

### Network analysis

Weighted Gene Co-Expression Network Analysis (WGCNA) was conducted using the Bioconductor R package *CWGCNA*. Mean signal intensity at replicate probes was calculated, as outlined in https://github.com/kobor-lab/EPICv2_QC_preprocessing/blob/main/EPICv2_DNAm_QC_preprocessing.Rmd. The relationship between methylation beta values and phenotype, in this case, HIV status or categorized total CTQ-SF scores, was assessed using the top 10 000 most variable probes. DNA methylation probe data were then compressed to genes using the *probestogenes* function. The WGCNA identifies modules of correlated gene expression and tests its association with the phenotypic variable. Finally, a causal WGCNA was conducted on the most significant module using the *diffwgcna* function. This enabled mediation models testing two causal directions: (1) a forward pathway (“module → module feature → phenotype”), where gene module differences precede the phenotype, and (2) a reverse pathway (“phenotype → module feature → module”), where module changes follow the phenotype, to identify potential causes and mediators [42].

### Statistical analysis

Data distribution was determined using a Shapiro-Wilk test. Continuous variables were analysed using the Wilcoxon Sum-Rank test. Medians and range are reported. Categorical variables were analysed using a chi-squared test.

## Results

### Participants

Participants without HIV were younger than those with HIV (p = 5.83e-08), with a median age of 26 years, compared to a median age of 33 years for the HIV group. For both groups, participants had a median education of 11 years. Women with HIV had a higher CTQ-SF total than the group without HIV (mean CM score 52.4 compared to 39.3) (Table 1).

**Table 1.**
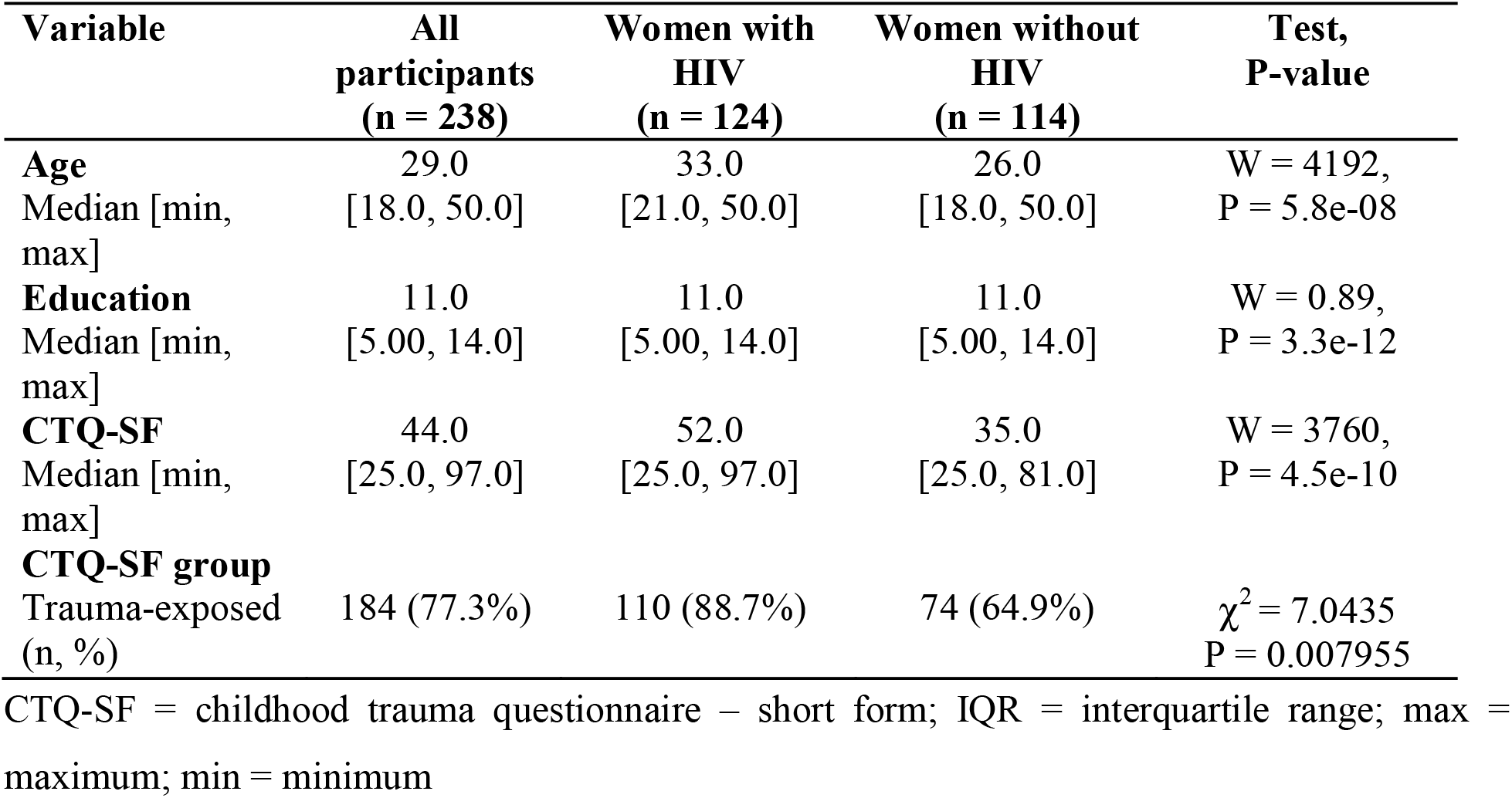
Demographic and clinical characteristics of participants with and without HIV.

### Site and regional DNA methylation profiles and associations with HIV

DNA methylation was not associated with HIV status at a site-specific level using the Benjamini-Hochberg method (pmin.obs. = 8.559e-07, FDR > 0.05), regardless of whether CTQ-SF scores were included as a covariate. An enrichment analysis of the top 100 CpG sites revealed genes in pathways involved in the major histocompatibility complex (MHC) protein complex and peptide antigen binding (Table 2). Gene ontology (GO) analyses identified that the plasma membrane cellular component contained the highest number of differentially methylated genes (DE = 9). However, given that 5536 genes are included under this category, other GO terms comprised a higher proportion of differentially methylated genes. The DMR analysis revealed no significant associations between DNA methylation and HIV status.

**Table 2.** Gene ontology for the top 100 CpG sites associated with HIV status.

| Gene ontology accession | Ontology | Term | N | DE | P.DE | FDR |
| --- | --- | --- | --- | --- | --- | --- |
| GO:0002396 | BP | MHC protein complex assembly | 19 | 2 | 7.30e-05 | 0.79 |
| GO:0002501 | BP | peptide antigen assembly with MHC protein complex | 19 | 2 | 7.30e-05 | 0.79 |
| GO:0042611 | CC | MHC protein complex | 23 | 2 | 1.05e-04 | 0.79 |
| GO:0098553 | CC | luminal side of endoplasmic reticulum membrane | 33 | 2 | 2.15e-04 | 0.10 |
| GO:0042605 | MF | peptide antigen binding | 33 | 2 | 2.23e-04 | 0.10 |
| GO:0002478 | BP | antigen processing and presentation of exogenous peptide antigen | 39 | 2 | 2.67e-04 | 0.10 |
| GO:0098576 | CC | luminal side of membrane | 44 | 2 | 3.64e-04 | 1.00 |
| GO:0019884 | BP | antigen processing and presentation of exogenous antigen | 48 | 2 | 3.67e-04 | 1.00 |
| GO:0003823 | MF | antigen binding | 64 | 2 | 6.30e-04 | 1.00 |
| GO:0012507 | CC | ER to Golgi transport vesicle membrane | 61 | 2 | 7.24e-04 | 1.00 |
| GO:0048002 | BP | antigen processing and presentation of peptide antigen | 70 | 2 | 8.01e-04 | 1.00 |
| GO:0005886 | CC | plasma membrane | 5536 | 9 | 1.14e-03 | 1.00 |
| GO:0150007 | BP | clathrin-dependent synaptic vesicle endocytosis | 2 | 1 | 1.37e-03 | 1.00 |
| **GO:2000567** | BP | regulation of memory T cell activation | 2 | 1 | 1.43e-03 | 1.00 |
| **GO:2000568** | BP | positive regulation of memory T cell activation | 2 | 1 | 1.43e-03 | 1.00 |
| **GO:0002485** | BP | antigen processing and presentation of endogenous peptide antigen via MHC class I via ER pathway, TAP-dependent | 2 | 1 | 1.48e-03 | 1.00 |
| **GO:0002419** | BP | T cell mediated cytotoxicity directed against tumor cell target | 3 | 1 | 1.53e-03 | 1.00 |
| **GO:0030134** | CC | COPII-coated ER to Golgi transport vesicle | 92 | 2 | 1.60e-03 | 1.00 |
| **GO:2000566** | BP | positive regulation of CD8-positive, alpha-beta T cell proliferation | 3 | 1 | 1.80e-03 | 1.00 |
| **GO:0062061** | MF | TAP complex binding | 3 | 1 | 1.86e-03 | 1.00 |

BP = biological pathway; CC = cellular component; MF = molecular function, DE = differentially expressed (number of genes represented by significant CpGs), P.DE = p value associated with differentially expressed genes, FDR = false discovery rate

### DNA methylation network profiles associated with HIV

Three of the four modules identified via WGCNA (Fig. 1A) showed significant differential methylation in relation to HIV status (Fig. 1B). In module three (ME3), the top-most differential WGCNA module, 51 out of 552 genes were upregulated in participants with HIV compared to without HIV (Fig. 1C). Mediation analysis for causal inference on ME3 and its features (Fig. 1D) identified 42 genes mediating the causal direction of HIV status → module gene → module, suggesting that HIV status drives differential DNA methylation in these genes (Fig. 1D).

**Figure 1.**
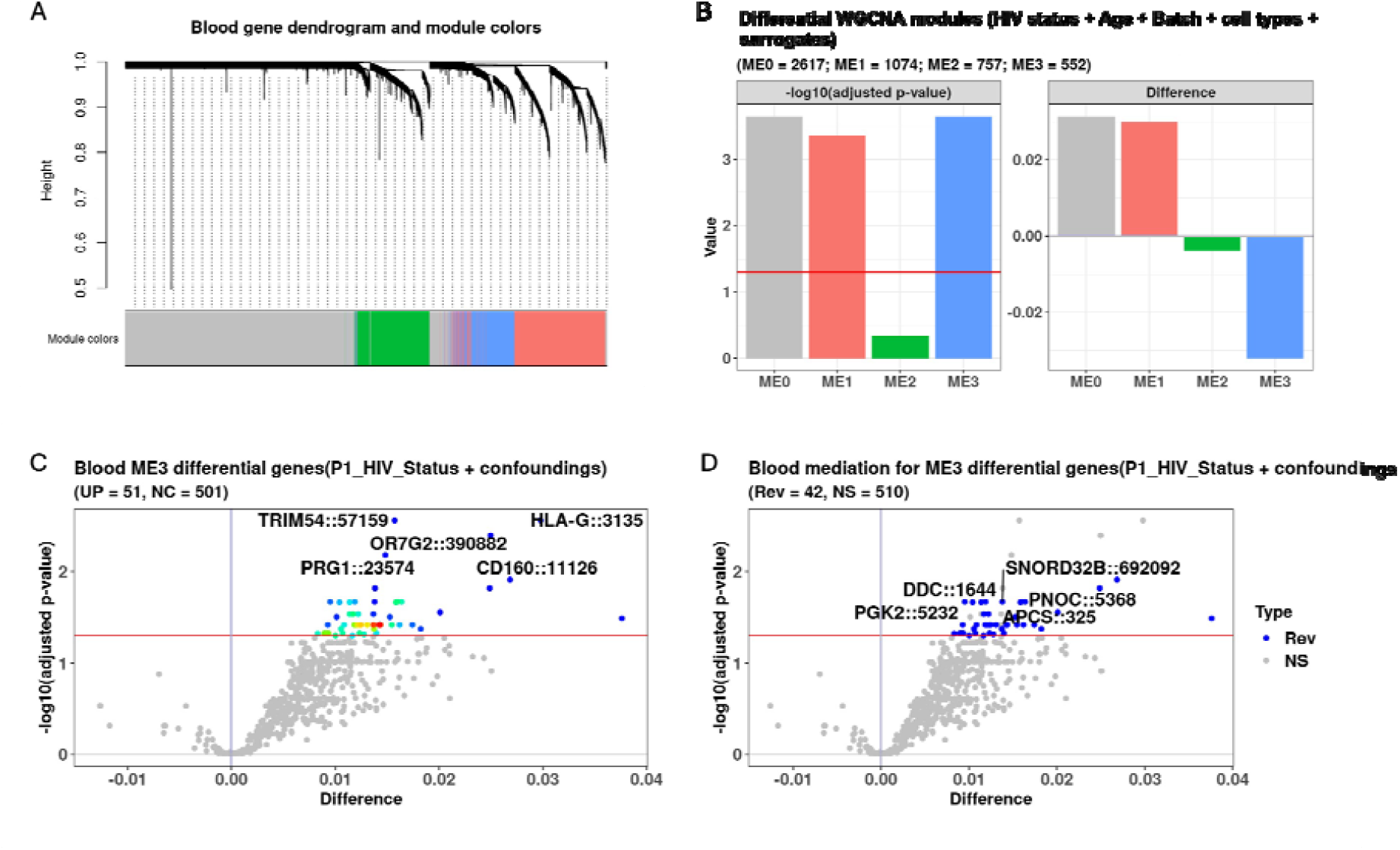
Four modules were significantly associated with HIV in WGCNA analyses. (A-B) Four modules were identified on WCGNA analysis. When adjusting for age, batch, cell type proportions and surrogate variables, three modules had significantly differential eigengene between HIV status groups, with the red line representing the significance threshold. Differentially methylated genes (C) and casual analysis (D) of module 3 identified 51 of 552 genes that were upregulated in PWH compared to those without HIV. Blue dots show gene mediating the causal forward pathway of HIV status → module gene → module.

### Regional DNA methylation profiles and CM associations

DNA methylation at CpG sites was not associated with CTQ-SF using the Benjamini-Hochberg method (pmin.obs = 2.0424e-06, FDR > 0.05). Enrichment analysis of the top 100 CpG sites revealed genes in pathways involved in transmembrane receptor proteins and tyrosine phosphatase activity, along with pathways involved in dendritic spine organization and development, and neuron projection organization (Table 3). Pathways involving nitrogen compound transport and the establishment of protein localization had the highest number of differentially methylated genes (DE = 2) (Table 3). Similar to the CpG association test result, th DMR analysis returned no statistically significant CM effects.

**Table 3.** Gene ontology for the top 100 CpG sites associated with childhood maltreatment.

| Gene<br>ontology<br>accession | Ontology | Term | N | DE | P.DE | FDR |
| --- | --- | --- | --- | --- | --- | --- |
| GO:0005001 | MF | transmembrane receptor protein<br>tyrosine phosphatase activity | 17 | 1 | 0.003 | 1 |
| GO:0019198 | MF | transmembrane receptor protein<br>phosphatase activity | 17 | 1 | 0.003 | 1 |
| GO:0101003 | CC | ficolin-1-rich granule membrane | 61 | 1 | 0.005 | 1 |
| GO:0150052 | BP | regulation of postsynapse assembly | 44 | 1 | 0.007 | 1 |
| GO:0015031 | BP | protein transport | 1422 | 2 | 0.007 | 1 |
| GO:0061001 | BP | regulation of dendritic spine<br>morphogenesis | 45 | 1 | 0.008 | 1 |
| GO:0035773 | BP | insulin secretion involved in<br>cellular response to glucose<br>stimulus | 67 | 1 | 0.009 | 1 |
| GO:0045184 | BP | establishment of protein<br>localization | 1704 | 2 | 0.010 | 1 |
| GO:0060997 | BP | dendritic spine morphogenesis | 60 | 1 | 0.010 | 1 |
| GO:0071705 | BP | nitrogen compound transport | 1901 | 2 | 0.012 | 1 |
| GO:0045202 | CC | synapse | 1656 | 2 | 0.013 | 1 |
| GO:0070820 | CC | tertiary granule | 164 | 1 | 0.013 | 1 |
| GO:0004725 | MF | protein tyrosine phosphatase<br>activity | 97 | 1 | 0.013 | 1 |
| GO:0097061 | BP | dendritic spine organization | 85 | 1 | 0.014 | 1 |
| GO:0099068 | BP | postsynapse assembly | 87 | 1 | 0.014 | 1 |
| GO:0106027 | BP | neuron projection organization | 95 | 1 | 0.015 | 1 |
| GO:0060996 | BP | dendritic spine development | 99 | 1 | 0.016 | 1 |
| GO:0001650 | CC | fibrillar center | 150 | 1 | 0.018 | 1 |
| GO:0071333 | BP | cellular response to glucose<br>stimulus | 147 | 1 | 0.018 | 1 |
| GO:0101002 | CC | ficolin-1-rich granule | 185 | 1 | 0.018 | 1 |

BP = biological pathway; CC = cellular component; MF = molecular function

### DNA methylation network profiles associated with CM

WGCNA identified one differentially methylated module associated with CM severity groups (Fig. 2A). In the module, 34 genes were upregulated and 3 genes were downregulated out of a total of 1074 genes (Fig. 2B). Mediation analysis for causal inference on the module and its features indicated that 17 genes mediated the causal forward pathway CM → module gene → module, suggesting that CM preceded the observed epigenetic effects (Fig. 2C).

**Figure 2.**
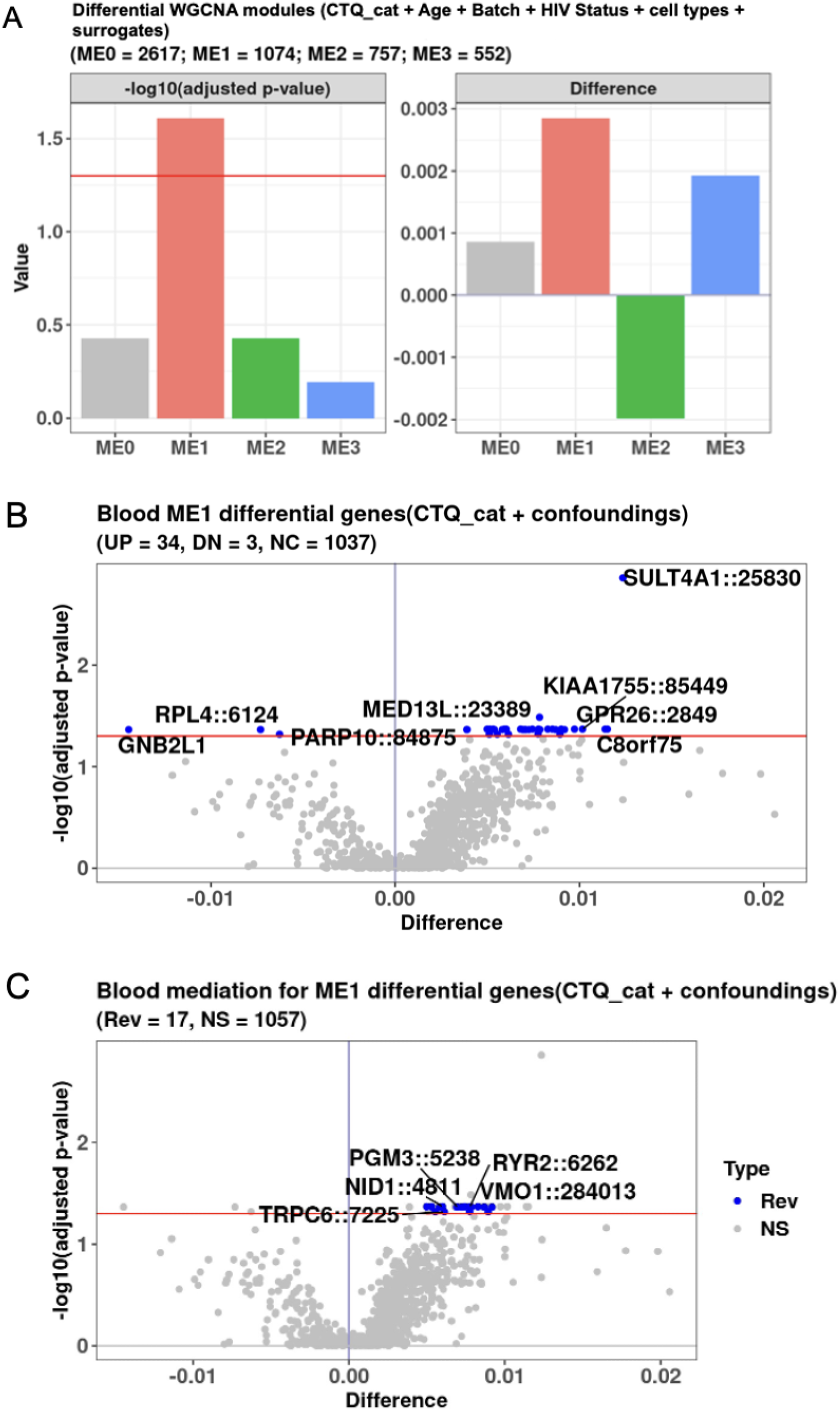
One module was significantly associated with CTQ score category in WGCNA analyses. (A) One DNA methylation module contained eigengenes significantly associated with CM severity group when adjusting for age, batch, cell type proportions, and surrogate variables. (B-C) Analysis of the module found 34 upregulated genes and 3 downregulated genes out of 1074 genes in those with at least mild-to-moderate compared to none-to-minimal CM severity. Blue dots indicate the 17 genes mediating the causal direction of childhood trauma → module gene → module.

## Discussion

This study aimed to identify DNA methylation signatures associated with HIV and CM in South African women. Identifying these methylation patterns may improve understanding of the epigenetic mechanisms underlying the long-term biological effects of HIV and CM, as well as their potential combined impact. Neither HIV status nor CM were associated with DNA methylation at site-specific or regional levels. However, network-level effects were found, with three out of four network modules differentially methylated in HIV, and one network module associated with CM. Both causal analyses inferred that HIV and CM preceded epigenetic effects.

In contrast to other published literature in which associate HIV-related DNA methylation with accelerated aging and abnormalities in immune-related gene expression [24,25,43,44], this study did not identify any significant DNA methylation regional patterns associated with HIV. Although no specific CpG site was significantly associated with HIV status, cg05671107_BC21 had the strongest association. This CpG is found in the promotor of the selenoprotein T-encoding *SELENOT* gene. Selenoprotein T plays a role in in protecting cells from oxidative stress, with reported neuroprotective effects on dopaminergic neurons [45]. Suboptimal functioning of selenoprotein T may therefore be related to neuropathological outcomes seen in PWH. Interestingly, studies report a negative correlation between serum selenium status and mortality in HIV-1 patients, with selenium supplementation being clinically beneficial [46–49].

Our methylation analyses consistently implicate the immune system in HIV and extend the existing literature by using DNA methylation approaches to further characterize immune-regulatory gene expression changes associated with HIV [22–27]. The top 3 biological pathways associated with HIV status were the MHC complex, peptide antigen assembly with MHC protein complex, and antigen processing and presentation of exogenous peptide antigen, all of which relate to the immune response.

WGCNA analysis identified 51 genes upregulated in HIV. Two of the top five upregulated genes are involved in immune functioning, *HLA-G* (Human Leukocyte Antigen-G) and *CD160*. HLA-G is an MHC molecule that inhibits allogeneic proliferation of CD4+ T, natural killer, and CD8+ T cell cytotoxicity, as well as the maturation of dendritic cells and activation of B cells. Inhibition of immune cells may be an indicator of an ineffective immune response. Less is known about the potential links between HIV and the three genes comprising the top five of the upregulated gene list. *TRIM54* (tripartite motif containing 54) regulates titin kinase and microtubule dependent signal pathways in striated muscles [50], *PRG1* (p53-responsive gene 1) is implicated in the inactivation of the p53 tumour suppressor protein in cancers [51], and *OR7G2* is responsible for the recognition and transduction of odorant signals [52].

CM was not associated with DNA methylation at CpG sites, and the functional relevance of the top five CpG sites is unclear as they do not map to the promotor regions of known genes. No significant CM–DMR associations were found, but the top three biological pathways associated with CM are involved in the regulation of post-synapse assembly, protein transport, and regulation of dendritic spine morphogenesis. This suggests that CM exerts its neurobiological effects on the developing central nervous system, as previously suggested [10].

WGCNA identified 34 genes upregulated in participants with at least minimal CM exposure. Of the top four genes related to brain function, sulfotransferase family 4A member 1 (*SULT4A1)* encodes a central nervous system-specific sulfotransferase involved in the metabolism of neurotransmitters [53], and polymorphisms of this gene have been associated with schizophrenia [54]. Mediator complex subunit 13L (MED13L) encodes a subunit of the Mediator complex [55], and is involved in early neurodevelopment [56]. G-protein coupled receptor 26 (*GPR26)* encodes the G-protein-coupled receptor, which is involved in cellular responses to environmental stimuli, neurotransmitters, and hormones, with sub-optimal function being implicated in neurodegenerative disorders [57]. An epigenetic study also demonstrated that silencing of this gene was associated with gliomas [58]. CM was also associated with ribosome biology in the current study. Poly (ADP-ribose) polymerase 10 (*PARP10*), involved in ribosomal function, was among the top four upregulated genes. In contrast, ribosomal protein L4 (*RPL4),* which encodes a ribosomal protein component of the cytoplasmic 60S subunit, was one of the top downregulated genes [59].

Although our site-and region-specific DNA methylation profiles cannot establish directionality, our network models suggest that HIV and CM are more likely associated with interconnected methylation profiles across multiple genes rather than isolated changes in single genes. Our results indicate an influence of HIV on immune pathways, and CM effects on neuronal pathways, particularly dendritic and synaptic structures, along with neurotransmitter function. Causal analysis also suggests that both HIV and CM exert effects on DNA methylation. Further research to understand the epigenetic mediation of HIV and CM on long-term health is needed.

### Limitations of the study

DNA is susceptible to several epigenetic modifications, including histone modifications and regulation by non-coding RNAs, with DNA methylation representing only one component. Accordingly, this study captures only a subset of the mechanisms underlying environmentally sensitive regulation of gene expression. Another limitation is the dynamic nature of DNA methylation, which varies across age, sex, and tissue or cell type. Epigenetic patterns vulnerable to environmental influence are not consistent across time or within specific cell type i.e. epigenetic differences that may have existed during childhood might not be present during adulthood. Our study tried to mitigate this by using established approaches, including only examining adult women and adjusting for age and cell type proportions. Cigarette smoking (nicotine use) could not be accounted for.

### Strengths of the study

A notable strength of this study is its consideration of DNA methylation as both a potential outcome and causal mediator of HIV and CM, allowing a closer examination of how epigenetic and environmental variables can interact with each other and potentially influence pathophysiology. Candidate gene findings are afflicted by non-reproducible false positives and associations are typically not robust enough to withstand genome-wide thresholds of significance [28]. In contrast, this study utilized a targeted panel consisting of more than 900 000 CpG sites with highly reproducible loci, thus offering a more complete picture of HIV- and CM-related methylation profiles across the genome.

## Conclusions

Using CpG site-specific, regional, network, and causal analysis, this study provides a more comprehensive understanding of the effects of HIV and CM on DNA methylation profiles. Immune-related DNA methylation pathways may serve as key candidates for elucidating the epigenetic effects of HIV. Likewise, the association between CM and neuronal processes warrants further investigation.

## Authors’ contributions

Conceptualization: SMJH; SL, SS; JSW; Data curation: AAR; GS; JSW; Formal analysis: AAR; Funding acquisition: SL; JSW; Investigation: AAR, JSW; Methodology: AAR, SMJH, GS, SL, SS, JSW; Project administration: GS, SS, JSW; Supervision: SMJH, GS, SS, JSW; Visualization: AAR ; Writing – original draft: AAR; Writing – review and editing: AAR, SMJH, GS, SL, SS, JSW

## Acknowledgements

We would like to acknowledge the participants, researchers and all staff involved in the parent study.

## Data Availability Statement

The data that support the findings of this study are available on reasonable request from the corresponding author. The data are not publicly available due to privacy or ethical restrictions.

